# Population scale whole genome sequencing provides novel insights into cardiometabolic health

**DOI:** 10.1101/2024.05.27.24307970

**Authors:** Yajie Zhao, Sam Lockhart, Jimmy Liu, Xihao Li, Adrian Cortes, Xing Hua, Eugene J. Gardner, Katherine A. Kentistou, Yancy Lo, Jonathan Davitte, David B. Savage, Carolyn Buser-Doepner, Ken K. Ong, Haoyu Zhang, Robert Scott, Stephen O’Rahilly, John R.B. Perry

**Author notes:** Correspondence to John R B Perry. denotes equal contribution (Yajie Zhao, Sam Lockhart, Jimmy Liu, Xihao Li, Haoyu Zhang, Robert Scott, Stephen O’Rahilly, John R.B. Perry).

## Abstract

In addition to its coverage of the non-coding genome, whole genome sequencing (WGS) may better capture the coding genome than exome sequencing. We sought to exploit this and identify novel rare, protein-coding variants associated with metabolic health in newly released WGS data (N=708,956) from the UK Biobank and All of Us studies. Identified genes highlight novel biological mechanisms, including protein truncating variants (PTVs) in the DNA double-strand break repair gene *RIF1* that have a substantial effect on body mass index (BMI, 2.66 kg/m^2^, s.e. 0.43, *P* = 3.7×10^-10^). *UBR3* is an intriguing example where PTVs independently increase BMI and type 2 diabetes (T2D) risk. Furthermore, PTVs in *IRS2* have a substantial effect on T2D (OR 6.4 [3.7-11.3], *P* = 9.9×10^-14^, 34% case prevalence among carriers) and were unexpectedly also associated with chronic kidney disease independent of diabetes status, suggesting an important role for IRS-2 in maintaining renal health. We identified genetic evidence of functional heterogeneity in *IRS1* and *IRS2*, suggesting a greater role for IRS-1 in mediating the growth promoting effects of insulin and IGF-I, while IRS-2 has a greater impact on glucose homeostasis likely through its actions in the pancreatic islet and insulin target tissues. Our study demonstrates that large-scale WGS provides novel mechanistic insights into human metabolic phenotypes through improved capture of coding sequences.

## Introduction

Genome-wide, hypothesis-free interrogation of the association between genomic variants and human traits and diseases in large populations has resulted in many key insights into the pathogenesis of common cardiometabolic disorders. The power of this approach has increased with the availability of population-scale whole exome sequencing (WES) data^1^. In contrast to earlier common variant genome-wide association studies (GWAS) where the majority of associated variants are non-coding^2–4^, and the causal gene is often unclear, studies leveraging rare protein-coding variation in gene-based collapsing tests more confidently identify causal genes and directions of effect relative to gene function. This approach more readily identifies novel causal pathways and mechanisms of disease for experimental interrogation^5^.

A recent advance has come from the widespread adoption of whole genome sequencing (WGS) in large population studies^6^. While the obvious advantage of WGS above WES is its ability to interrogate the non-coding genome, it has also been demonstrated that WGS identifies more functional coding variation than exome-sequencing based technologies^7^.

Here, we sought to leverage the increased sample size and purported enhanced capture of rare coding variation from UK Biobank WGS data^7^ to provide novel insight into the genetic basis of two cardiometabolic traits of major significance to population health; Type 2 Diabetes (T2D) and Body Mass Index (BMI). Previous large-scale WES studies have identified several genes harbouring rare protein coding variants of large effect for these traits^8–12^ including examples where heterozygous loss of function either increases (e.g. *GIGYF1* for T2D^13^, *BSN* for obesity^9,14^) or decreases the risk of disease (e.g. *MAP3K15* for T2D^12^, *GPR75* for obesity^8^). By extending these analyses to consider WGS data in more than 480K UK Biobank participants, we identify five novel associations which we replicate in 219,015 individuals from the All of Us study. These findings include T2D risk increasing PTVs in the gene *IRS2*, encoding a key node in the insulin/IGF-1 signalling cascade, which also increased risk of CKD independent of diabetes status, and PTVs in the ubiquitin-ligase UBR3 with independent effects on BMI and T2D risk. Together, these findings identify novel genetic determinants of cardiometabolic risk and highlight impaired IRS2-mediated signalling as an unexpected candidate mechanism of renal disease.

## Results

To identify genes associated with either adult BMI or T2D risk, we performed association testing using WGS data available in up to 489,941 UK Biobank participants (see methods). This represents a sample size increase of up to 71,505 individuals compared to our recent WES analyses of the same cohort^9,11^, attributable to both an increase in the number of sequenced samples (N= 35,725) and the inclusion of individuals of non-European ancestry (N= 64,609). Individual gene-burden tests were performed by collapsing rare (MAF < 0.1%) variants across 19,457 protein-coding genes. We tested three categories of variants based on their predicted functional impact: high-confidence protein-truncating variants (PTVs), and two overlapping missense masks that used a REVEL^15^ score threshold of 0.5 or 0.7. This yielded a total of 81,350 tests (40,750 tests for T2D and 40,600 tests for BMI) for gene masks with at least 30 informative rare allele carriers, corresponding to a conservative multiple-test corrected statistical significance threshold of *P* < 6.15 × 10^-7^ (0.05/81,350).

Genetic association testing identified a total of 21 genes with at least one mask associated at this threshold with adult BMI (n=10 genes) or T2D (n=12 genes). (Figure 1 and Supplementary Table 1) The only overlapping association between the two traits was with PTVs in *UBR3*. Our WGS analysis confirmed previously reported gene associations using WES for BMI including PTVs and damaging missense variants in *MC4R*, *UBR2*, *SLTM* and *PCSK1*, *BSN*, *APBA1* and *PTPRG*^8,10,13,14^. Our WGS analysis also confirmed previously reported gene associations using WES for T2D including PTVs in *GCK*, *HNF1A*, *GIGYF1* and *TNRC6B*, and missense variants with REVEL >= 0.7 in *IGF1R*^11,13^. For most of these genes, we observed stronger associations using WGS than we previously reported using WES, with an overall 29% increase in mean chi-square values for these associated genes using similar variant masks. (Supplementary Table 2) Our WGS gene-burden test appeared statistically well calibrated, as indicated by low exome-wide test statistic inflation (*λ*_GC_ = 1.15 for BMI and 1.20 for T2D) and by the absence of significant associations with any synonymous variant masks (included as a negative control). To replicate our findings in UK Biobank WGS data, we implemented an identical variants annotation workflow for genes identified from UK Biobank and run gene-burden testing using WGS data derived from 219,015 participants in the All of Us studies.

**Figure 1.**
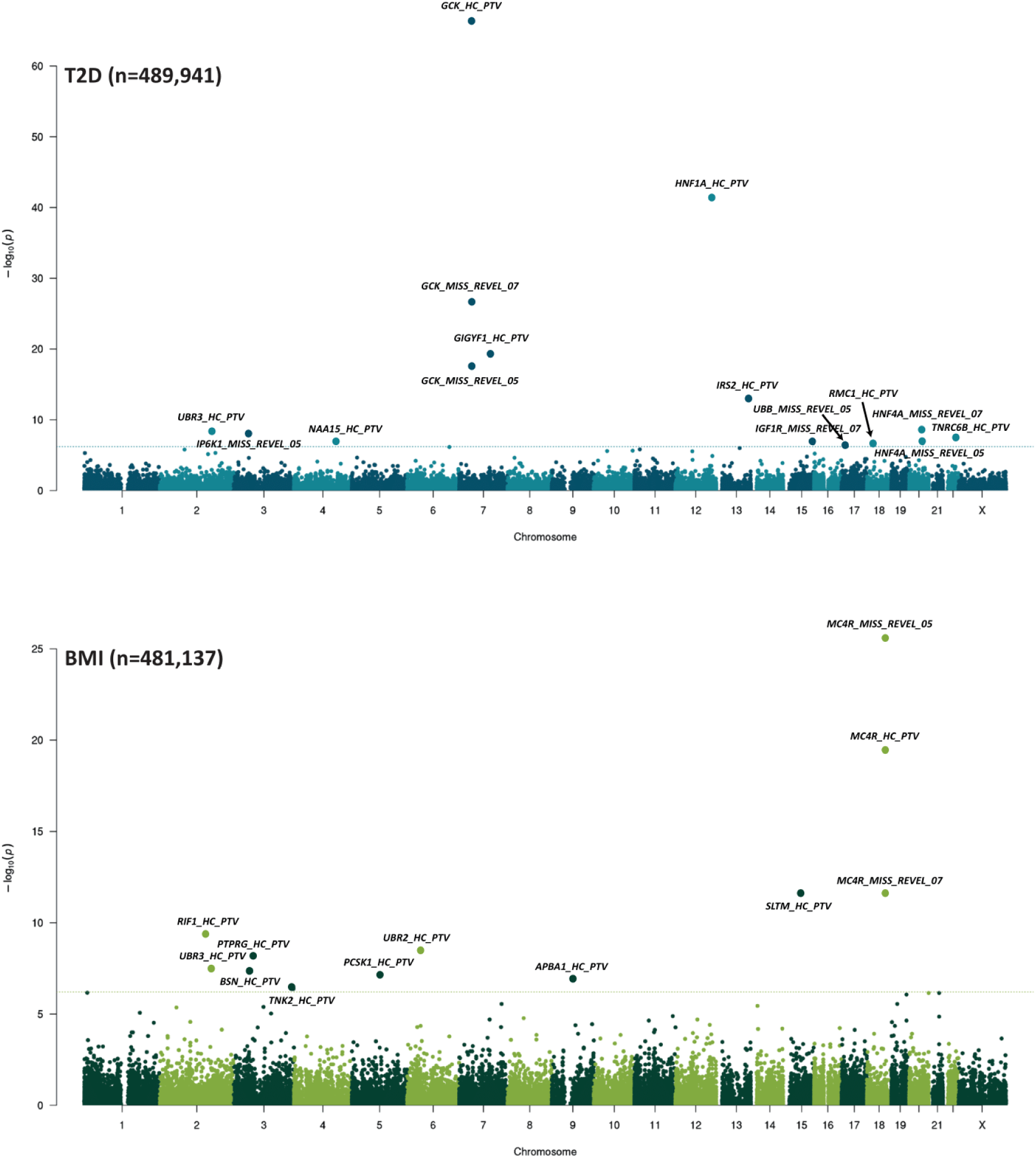
| Genome-wide multi-ancestry gene burden test for BMI (top panel) and T2D (bottom panel) in UK Biobank. Manhattan plots showing gene burden test results for BMI and T2D. Genes passing exome-wide significance (*P <* 6.15 × 10^-7^ (0.05/81,350)) are labelled. Points are annotated with variant predicted functional mask (MISS REVEL; missense variants with REVEL scores (above 0.5 or 0.7), HC PTV; high confidence protein truncating variants).

At the three genes that we newly identified for BMI, PTVs conferred higher adult BMI: *RIF1* (effect per allele = 2.66 kg/m^2^, s.e. = 0.43, *P* = 3.7 × 10^-10^, carrier *n* = 117) - encoding an effector in the non-homologous end-joining pathway activated in response to double stranded DNA-breaks^16^, *UBR3* (2.41 kg/m^2^, s.e. = 0.44, *P* = 3.6 × 10^-8^, carrier *n* = 111) - an E3-ubiquitin ligase that is highly expressed in sensory tissues^17^, and the non-receptor tyrosine kinase *TNK2* (0.88 kg/m^2^, s.e. = 0.17, *P* = 4.2 × 10^-7^, carrier *n* = 702). Two of these three novel gene associations with BMI were replicated in All of Us (at *P*<0.05, Figure 2 and Supplementary Table 3): *RIF1* (2.58 kg/m^2^, s.e. = 1.17, *P* = 2.8 × 10^-2^, carrier *n* = 39) and *UBR3* (3.21 kg/m^2^, s.e. = 0.84, *P* = 1.3 × 10^-4^, carrier *n* = 67). (Supplementary Table 4) Previous GWAS studies also identified loci associated with BMI and T2D within 500kb of *RIF1* (T2D: rs6567160:T, beta=0.018, s.e. = 0.003, *P* = 4.1 × 10^-10^) and *TNK2* (BMI: rs34801745:C, beta=0.013, s.e. = 0.002, *P* = 7 × 10^-11^; T2D: rs6800500:C, beta=0.03, s.e. = 0.003, *P* = 2.4 × 10^-21^). (Supplementary Table 5). Using a variant to gene mapping method^18^(see Methods), GWAS signals at both the *RIF1* and *TNK2* loci could be confidently linked to the function of these genes, e.g. we observed colocalisation between eQTLs for both RIF1 and TNK2, with decreased expression corresponding to increased BMI, directionally concordant with their rare PTV effects (Supplementary Table 5).

**Figure 2.**
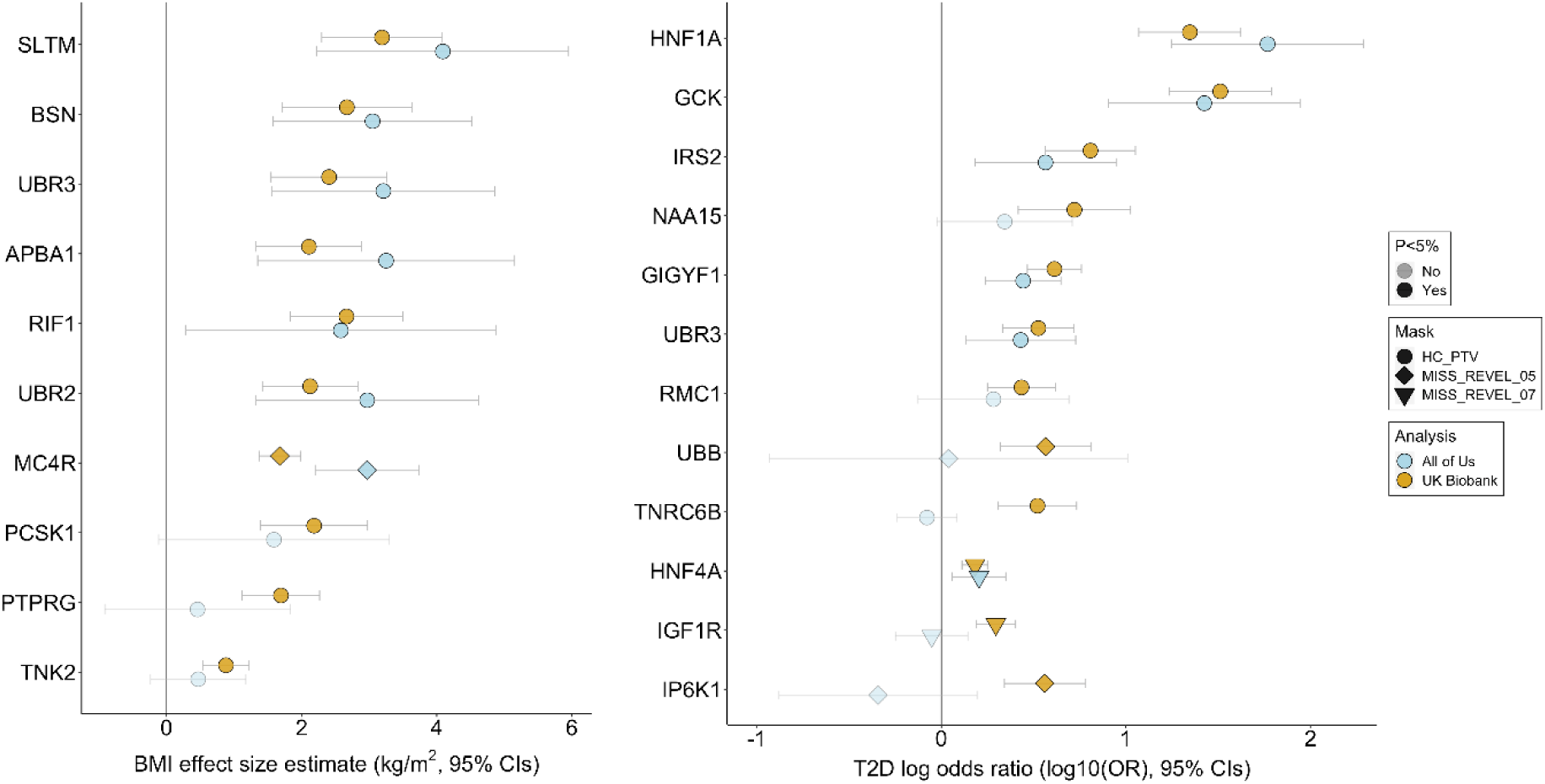
| Discovery and replication of significant associations with BMI (left) and T2D (right) in UK Biobank and All of Us study. For single genes with multiple significant associations, only the most significant association is displayed. Odds of T2D are plotted on a Log10-scale.

At the seven genes that have not been previously implicated via population-scale studies for T2D, PTVs conferred higher risk for T2D: *IRS2* (OR per allele =6.4, 95% CI [3.7-11.3], *P* = 9.9 × 10^-14^, carrier *n* = 58, 34% case prevalence among carriers) - encoding a key adaptor molecule in the insulin-signaling cascade, *UBR3* (OR =3.4, 95% CI [2.1-5.2], *P* = 6.1 × 10^-9^, carrier *n* = 115, 23% case prevalence) - encoding a component of N-terminal acetyltransferase complexes^19^, *NAA15* (OR =5.3, 95% CI [2.6-10.6], *P* = 1.2 × 10^-7^, carrier *n* = 39, 31% case prevalence) and *RMC1* (OR =2.7, 95% CI [1.8-4.2], *P* = 3.4 × 10^-7^, carrier *n* = 138, 20% case prevalence) - encoding part of a protein complex critical for lysosomal trafficking and autophagy^20,21^. (Supplementary Table 4) Our missense mask also identified associations with *IP6K1* (OR =3.6, 95% CI [2.2-6.0], *P* = 8.5 × 10^-9^, carrier *n* = 84, 26% case prevalence) - encoding inositol phosphokinase, the known MODY gene *HNF4A* (OR =1.5, 95% CI [1.3-1.8], *P* = 3.1 × 10^-9^, carrier *n* = 1,386, 13% case prevalence), and *UBB* (OR =3.7, 95% CI [2.1-6.4], *P* = 5.8 × 10^-7^, carrier *n* = 66, 26% case prevalence) - encoding ubiquitin. (Supplementary Table 4) Three of these seven gene associations with T2D were replicated in All of Us: *IRS2* (OR =3.7, 95% CI [1.5-8.9], *P* = 3.9 × 10^-3^, carrier *n* = 40, 30% case prevalence), *UBR3* (OR =2.7, 95% CI [1.3-5.3], *P* = 5 × 10^-3^, carrier *n* = 67, 25% case prevalence) and *HNF4A* (missense variants with REVEL >= 0.7: OR =1.6, 95% CI [1.2-2.2], *P* = 4.7 × 10^-3^, carrier *n* = 293, 20% case prevalence; missense variants with REVEL >= 0.5: OR =1.3, 95% CI [1.1-1.7], *P* = 1.5 × 10^-2^, carrier *n* = 634, 17% case prevalence). (Supplementary Table 4)

There were also common GWAS loci associated with BMI and T2D within 500kb from *IRS2* (T2D: rs9301365:T, beta=0.024, s.e. = 0.003, *P* = 2.1 × 10^-16^), *RMC1* (BMI: rs891387:T, beta=0.021, s.e. = 0.002, *P* = 9.3 × 10^-35^; T2D: rs1788819:G, beta= 0.032, s.e. = 0.003, *P* = 4 × 10^-21^), *IP6K1* (BMI: rs11713193:A, beta=0.025, s.e. = 0.002, *P* = 3 × 10^-48^; T2D: rs7613875:A, beta= 0.025, s.e. = 0.003, *P* = 4.8 × 10^-16^), *HNF4A* (BMI: rs2284265:T, beta=0.012, s.e. = 0.002, *P* = 4.8 × 10^-8^; T2D: rs12625671:C, beta= 0.067, s.e. = 0.004, *P* = 1.7 × 10^-68^) and *UBB* (BMI: rs1075901:C, beta=0.012, s.e. = 0.002, *P* = 4.4 × 10^-13^)(Supplementary Table 5). Of these, we could confidently link variants at the *IRS2* and *HNF4A* T2D loci with the corresponding gene’s function (Supplementary Table 5).

As a further sensitivity analyses, we performed ’leave-one-out analyses’ which confirmed that none of the above gene-level associations was driven by a single rare variant. (Supplementary Table 6) Furthermore, all novel associations exhibited similar effects in published results using WES data from UK Biobank but at sub-threshold significance (*P* <= 8.3 × 10^-5^).

To assess its added value, we compared our all-ancestries based approach to an European-only analysis using otherwise identical analytical parameters. Among the 27 significant associations we identified, 21 had a stronger *P*-value in the all-ancestries analysis with an overall 4.6% increase in mean chi-square values. To similarly quantify the gain in statistical power using WGS, in the UK Biobank sample with both WGS and WES data available we compared the gene-burden test statistics from WGS and WES for the genes identified in our discovery analysis. On average, we observed a 21% increase in chi-square in the WGS analysis (Supplementary Table 2). Among the 26 gene-masks we compared, WGS data included (median, IQR: 12, 4-16) more variants compared to WES. Moreover, sensitivity gene-burden tests considering only those additional carriers identified by WGS (i.e. not identified by WES data), 16 of the 23 gene-mask combinations with at least 5 carriers showed a nominally significant association (*P* < 0.05) with the target phenotype, indicating that additional coding variants identified by WGS are likely to be functionally relevant. In contrast, some variants were identified by WES-only. Gene masks of these variants (with at least 5 carriers) showed not even nominal significant association with the target phenotype. Our findings confirm and quantify the enhanced coverage of coding variants provided by WGS above WES in UK Biobank.

### A phenotypic association scan of identified genes reveals a novel role for *IRS2* **in human kidney health**

To explore the broader phenotypic effects of our identified BMI-raising and T2D risk genes we conducted a phenotypic association scan for each gene variant mask significantly associated with T2D and BMI in our discovery analysis. (Supplementary Table 7 and 8). We observed several expected associations, for example between T2D risk genes with HbA1c and glucose and between BMI genes with whole body fat mass (Supplementary Figure 3). However, we were intrigued to observe a novel, highly statistically significant association of *IRS2* PTVs with lower Cystatin-C-derived estimated glomerular filtration rate (eGFR, effect = -12.92 mL/min/1.73m^2^, s.e. = 1.87, *P* = 4.9 × 10^-12^, carrier *n* = 55). This effect of *IRS2* PTVs on renal function was consistently observed across three different methods of GFR estimation (Figure 4).

This association does not simply reflect the consequences of T2D-mediated chronic hyperglycaemia on renal function as it was also observed in carriers of PTVs in *IRS2* without a diagnosis of T2D (Cystatin-C-derived eGFR: effect = -10.42 mL/min/1.73m^2^, s.e. = 2.24, *P* = 3.3 × 10^-6^, carrier *n* = 36). Consistent with a renoprotective role for IRS-2 in humans, PTVs in *IRS2* were associated with a ∼4-fold increase in odds of chronic kidney disease (CKD) (OR =4.0, 95% CI [1.9-8.6] *P* = 3.1 × 10^-4^, carrier *n* = 58, 14% case prevalence, Figure 4). These results identify *IRS2* as a T2D risk gene with an independent effect on CKD risk.

We also observed that PTVs in the adaptor protein *GIGYF1* conferred beneficial effects on serum lipids, consistent with previous findings^22^, but deleterious effects on renal function including a ∼2-fold increase in odds of CKD (Supplementary Table 7 and Figure 3). We also note a striking reduction in circulating SHBG (sex hormone binding globulin) levels in carriers of predicted damaging missense mutations in *HNF4A* (effect = -6.4 nmol/L, s.e. = 0.73, *P* = 7.5 × 10^-19^, carrier *n* = 1200), which has been reported to regulate *SHBG* transcription *in vitro*^23^. PTVs in *RMC1* conferred higher triglycerides, lower HDL (and therefore higher TG:HDL ratio) and increased risk of algorithmically defined MAFLD, a pattern of association suggestive of lipotoxic insulin resistance.

**Figure 3.**
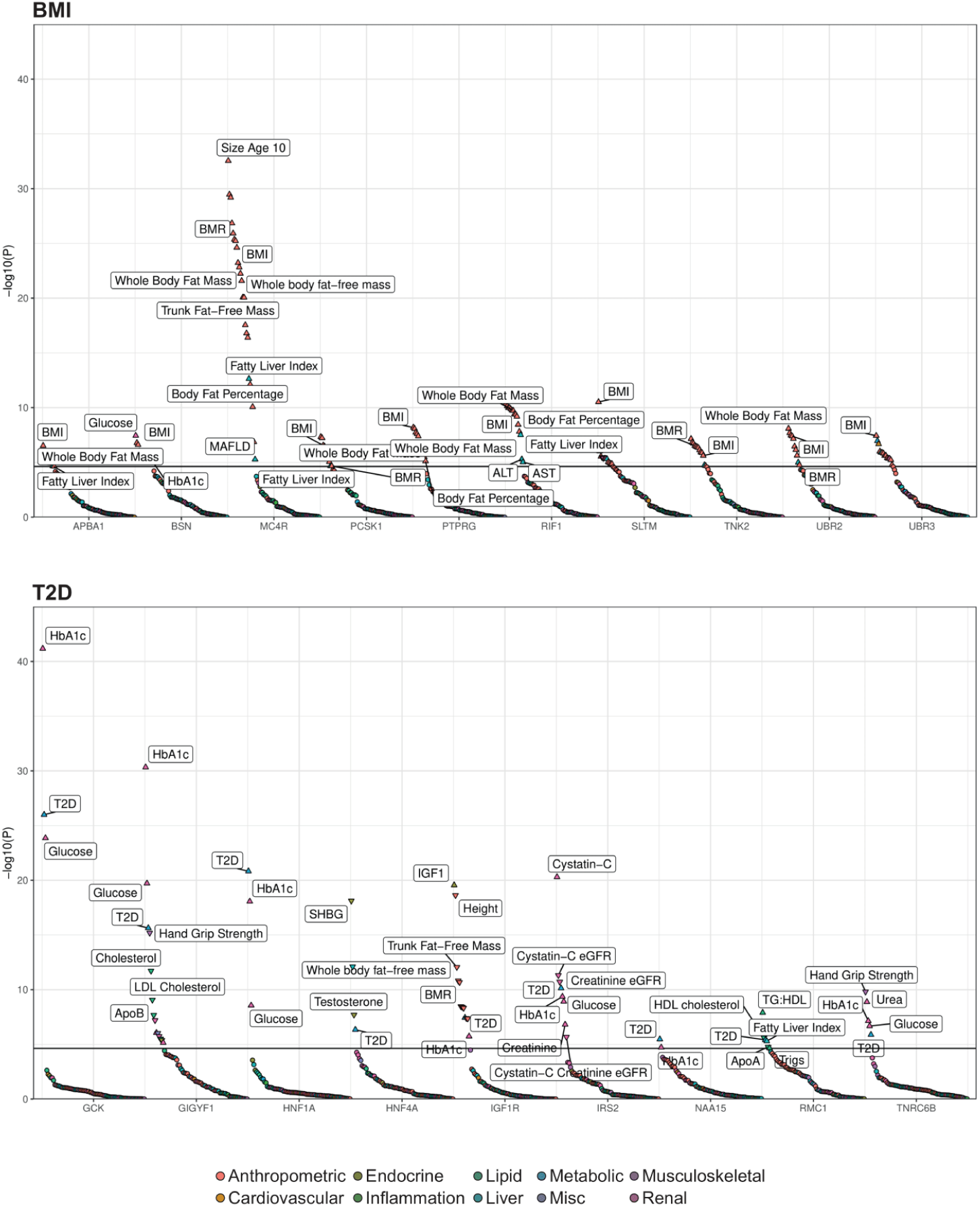
| Phenotypic association scans of BMI (top) and T2D (bottom) associated genes in UK Biobank. The **most** significant Gene x Mask association with BMI or T2D was assessed on a panel of 79 traits (see methods). Dots are coloured according to classification of phenotype; the orientation of triangles indicate the direction of effect for significant traits. For clarity, only a subset of traits and the most significant Gene x Mask association (for genes with >1 mask significantly associated with T2D or BMI) are displayed. *UBR3*, which was associated with both T2D and BMI in our discovery analysis is presented alongside BMI risk genes only to avoid duplication. The solid horizontal lines represent a Bonferroni-corrected threshold for statistical significance of 2.35 x 10^-5^ (0.05/2132 Phenotype x Mask associations).

**Figure 4.**
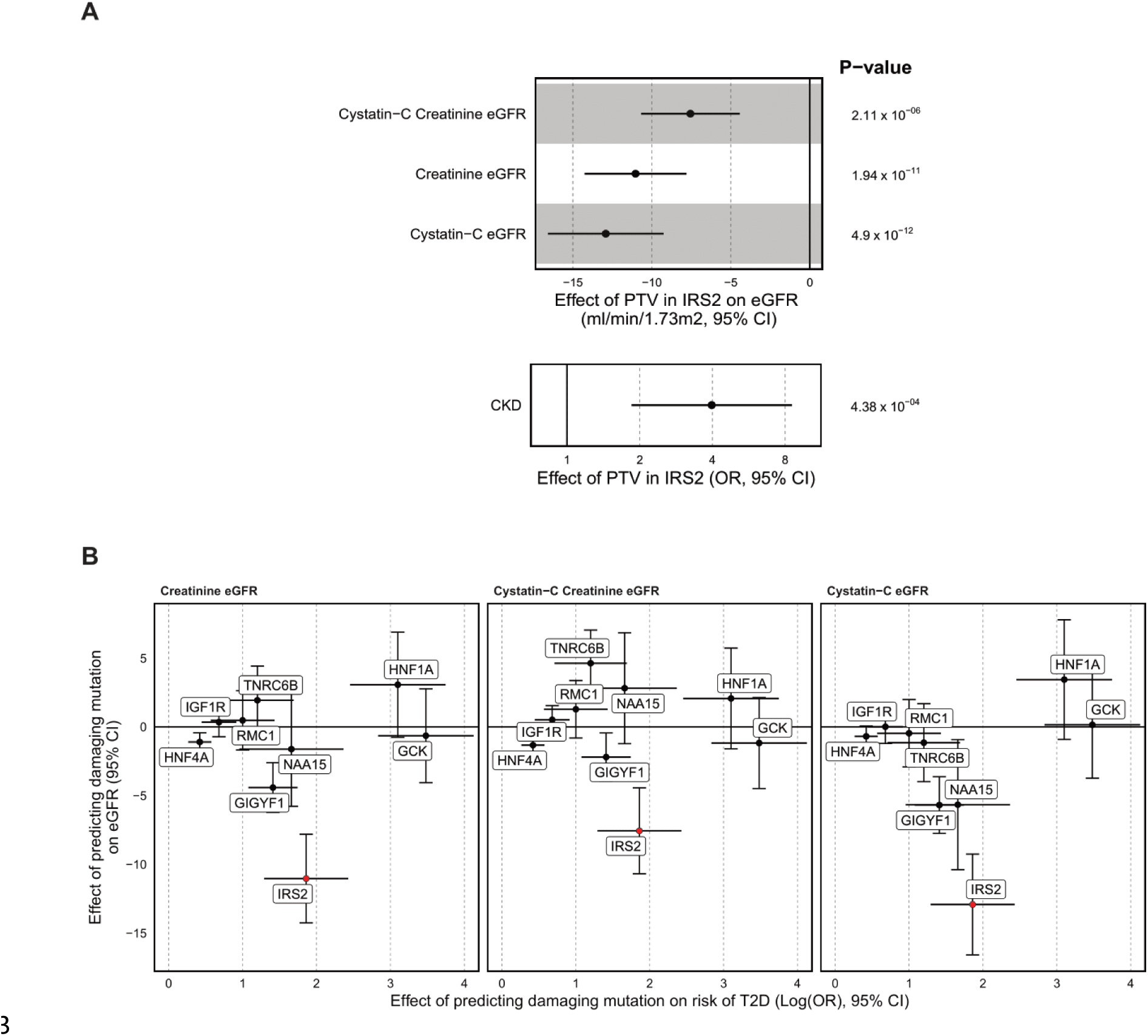
| Loss of function mutations in *IRS2* increase CKD risk. **A:** The effect of protein truncating variants in *IRS2* on various measures of eGFR (ml/min/1.73m^2^) and algorithmically defined CKD (OR) and P-values from linear and logistic regression, respectively are illustrated. Odds of CKD are plotted on log-scale. **B:** The effect of rare predicted damaging mutations in the labelled genes on T2D risk are plotted against the effect on eGFR (across three different methods of estimation) to illustrate that the effect of PTVs in *IRS2* on renal function appear independent of its effect on T2D. For clarity, only the gene x mask combination most significantly associated with T2D is plotted.

### Evidence of functional diversity in IRS1/IRS2 mediated signalling

IRS-1 and IRS-2 are critical nodes in the insulin/IGF-1 signalling cascade. They are recruited to and phosphorylated by the activated insulin receptor, serving as essential adaptor molecules to mediate downstream signalling. An interesting finding from mouse genetic studies is that *IRS1*-knockout mice do not show fasting hyperglycaemia, despite evidence of insulin resistance and reduced body size, consistent with impaired growth due to IGF-1-resistance^24,25^. In contrast, IRS2-knockout mice are comparable in size to their littermate controls but exhibit fasting hyperglycaemia and glucose intolerance due to failed beta-cell compensation^26^. To determine if similar phenotypic heterogeneity is present in humans, we compared the effects of *IRS1* and *IRS2* loss of function. (Supplementary Table 7 and 8) Consistent with the described mouse biology, human carriers of PTVs in *IRS1* had reduced fat-free mass and reduced height – suggestive of impairment in the anabolic effects of IGF-1 signalling. In contrast, carriers of PTVs in *IRS2* had no changes in lean mass or height, but a substantially increased risk of T2D (Figure 5). These findings suggest that the functional specificity of IRS-1/IRS-2 previously described in mice is conserved in humans; IRS-1 likely mediates the effects of IGF-1 signalling on linear growth and lean mass, whereas IRS-2 is relatively more important for glucose tolerance, likely due to its key regulatory actions in the pancreatic beta cell.

**Figure 5.**
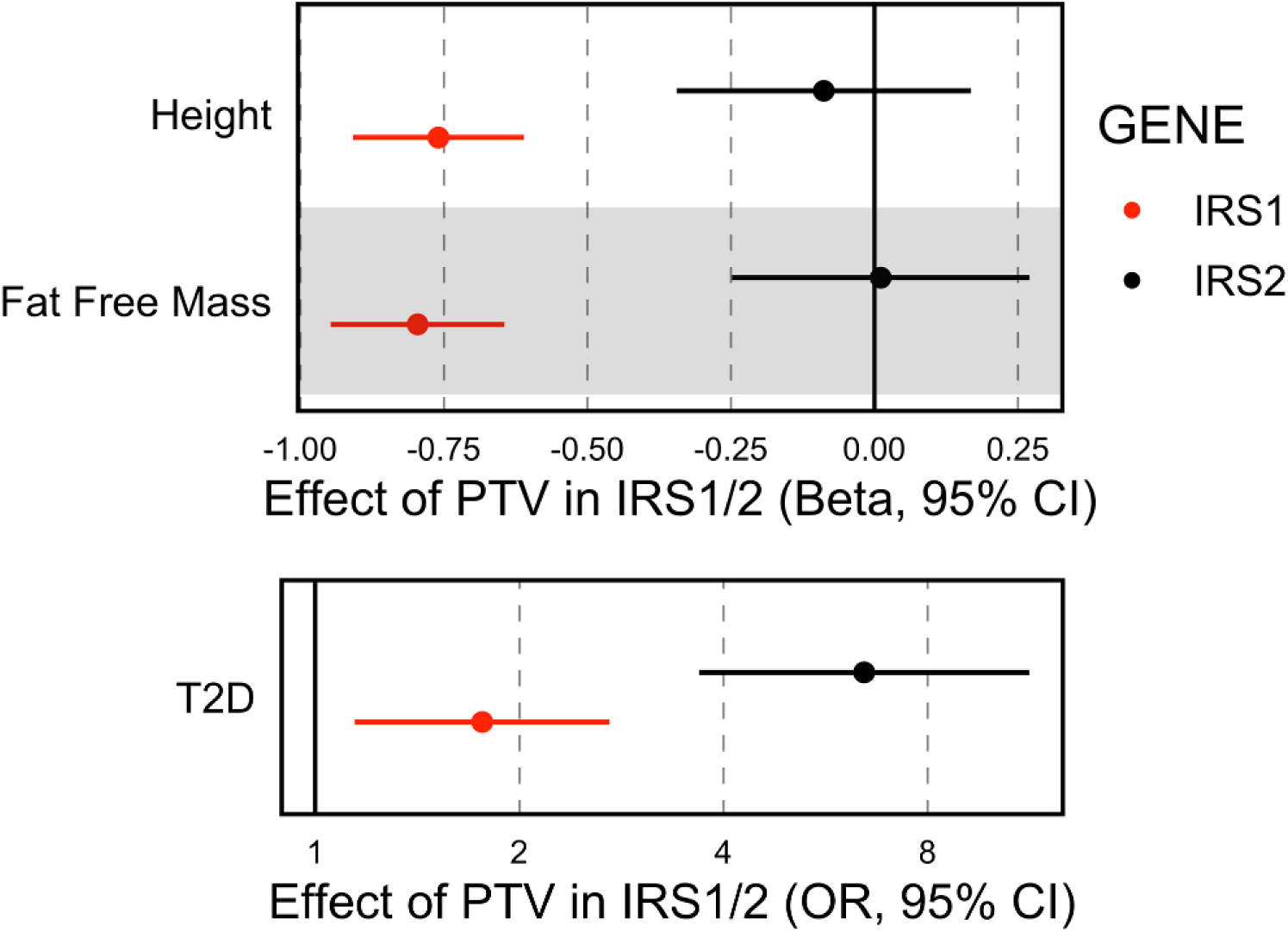
| Genetic evidence for functional heterogeneity of insulin receptor substrates in humans. Studies in mice have demonstrated that loss of IRS-1 markedly reduces body size with a modest effect on blood glucose, whereas loss of IRS-2 causes severe hyperglycaemia without affecting body size. We identify consistently divergent effects of PTVs in *IRS1* and *IRS2* in humans. Effect of PTVs in *IRS1* and *IRS2* on continuous traits are plotted as standardised betas and as odds ratio for T2D. Odds of T2D are plotted on a Log-scale.

### E3-ubiquitin ligases UBR2 and UBR3, body composition and type 2 diabetes risk

*UBR2* and *UBR3* are related E3-ubiquitin ligases. *UBR2* is a canonical N-recognin which recognises modified N-terminal amino acid residues (so-called N-degrons) and ubiquitinates these proteins to target them for degradation^27,28^. *UBR3* shares weak homology to *UBR2* and while it contains a UBR-domain which mediates recognition of modified amino acids by *UBR2*, it does not possess N-recognin activity but does mediate N-terminal ubiquitination via an as of yet unknown degradation signal^27,29^. In our discovery analyses, *UBR2* and *UBR3* PTVs were both associated with increased BMI, but only *UBR3* conferred a significant increase in T2D risk (Figure 1 and 3), consistent with distinct molecular actions of these proteins.

Importantly, the association of PTV in *UBR3* and T2D was not solely due to increased BMI as the effect on T2D was only partially attenuated after adjustment for BMI (OR =2.5, 95% CI [1.5-4.1], *P* = 2.7 × 10^-4^). To gain further insight into the mechanism through which UBR3 disruption increases T2D risk, we examined associations with body fat distribution and surrogate markers of insulin resistance measured in UKBB, SHBG and TG:HDL (Supplementary Table 8). We found no evidence for an effect of PTVs in *UBR3* on body fat distribution as assessed by WHRadjBMI and inconsistent effects on the surrogate markers of insulin resistance TG:HDL, which was not regulated, and SHBG which was nominally decreased.

Interestingly, UBR2 has been implicated in regulation of muscle mass in several mouse studies^30–32^. Therefore, we assessed the effect of *UBR2* and *UBR3* PTVs on lean and fat mass measured by bioimpedance in UK Biobank. Carriage of a PTV in *UBR2* or *UBR3* conferred higher whole body fat mass and, while *UBR2* PTV carriers showed a nominal increase in whole body fat free mass, this association was modest and likely to be a secondary effect of increased adiposity (Supplementary Table 8).

While we did not observe any notable effects of *UBR3* PTVs on fat-free mass measurements, maximum hand-grip strength was nominally increased (Supplementary Table 8).

## Discussion

By conducting the first genome-wide multi-ancestry gene burden test using WGS data from a cumulative total of >700,000 individuals, we identified several novel BMI and T2D-associated genes. Compared with previous European-only analysis based on WES data, we increased carrier number and statistical power by incorporating all individuals with available WGS data. Importantly, we demonstrate that our findings from UK Biobank are robust and reproducible, as several were replicated in an independent US population-based study (All of Us) which has considerably different demographics, notably its younger age, higher baseline prevalence of T2D and enhanced ethnic diversity^33^.

Our study also highlights some emerging challenges in conducting rare-variant association studies across diverse populations. For example, we failed to replicate two gene masks using All of Us data – *GCK* (Missense variants, REVEL>=0.5) and *IGF1R* (REVEL>=0.7). Both genes are robustly associated with T2D in UK Biobank, have >100 informative carriers in the All of Us cohort and have a high probability of being true based on either known clinical associations with T2D (*GCK*) or orthogonal support from common variant association studies (*IGF1R*)^11^. This may reflect specific challenges in the fidelity of missense classification tools across different pools of rare missense variants present in different cohorts with varying ethnic composition.

Our results provide several novel biological insights into the determinants of human cardiometabolic health. The association with *RIF1*, a gene implicated in telomere regulation, DNA repair and replication timing, expands the list of DNA damage response genes involved in metabolic health^10^. The biological mechanisms behind these associations remain unclear. However, as the same variants in *RIF1* showed not even nominal association with recalled childhood adiposity in UK Biobank (*P*>0.05), contrasting with their robust association with adult BMI (*P* = 3.7 × 10^-10^), we speculate that mechanisms that regulate neuronal degeneration might influence risk of adult-onset obesity^9^.

We identified a robust, replicable association of PTVs in the critical signalling node in the insulin/IGF1-pathway, IRS-2, and T2D with carriers exhibiting >3.6-fold increase in odds of diagnosis with T2D (OR 3.68 in All of US and 6.45 in UK Biobank). While insulin resistance is well known as a necessary antecedent to the development of T2D, there is a longstanding interest in discerning the role of specific nodes in the insulin signalling cascade in the development of insulin resistance and its related complications. By necessity, this work has largely been done in animal models and the translational relevance of these findings to human health is uncertain. Candidate-gene testing and exome-sequencing studies of probands with extreme phenotypes identified in the clinical setting have been leveraged to provide insight into the function of several nodes of the insulin-signalling cascade in humans (*INSR*, *PIK3R1* and *AKT2*) ^34–39^, but our findings place *IRS2* as the first component of the insulin signalling cascade to be definitively linked to T2D via study of rare LOF variants using an unbiased population-based sequencing approach.

To gain further insight into the specific phenotypic consequences of insulin/IGF1 resistance mediated by *IRS2* PTVs, we compared carriers of these variants with carriers of the other broadly expressed IRS protein, IRS-1. We found that PTVs in *IRS1* conferred a much more modest effect on T2D risk compared to *IRS2*, but significantly reduced height and lean mass, phenotypes which were not associated with *IRS2* PTVs. These findings recapitulate observations first made in lower organisms – IRS1 knockout mice are insulin resistant and small but develop only modest dysglycaemia due to compensatory changes in the beta-cell^24,25^. In contrast, *IRS2* knockout mice grow normally, and exhibit significant hepatic and skeletal muscle insulin resistance, but in contrast to their *IRS1* counterparts develop severe dysglycaemia due to beta-cell failure^26^. Our results suggest that functional heterogeneity in IRS-1/IRS-2-mediated insulin/IGF1-signaling is conserved across species and is consistent with an important function of IRS-2 in human beta-cell health, as has been demonstrated in mice^40^. Future recall by genotype studies of *IRS2* PTV carriers with detailed assessment of glucose homeostasis and insulin sensitivity will be key in determining the relative contribution of insulin resistance and beta-cell failure to the development of T2D in *IRS2*-haploinsufficiency.

To gain a broader perspective of the effects of T2D risk and BMI-raising genes we conducted a phenotypic association scan for these genes. Remarkably, we observed that PTVs in *IRS2* significantly reduce eGFR independent of T2D status and cause a 4-fold increase in the odds of algorithmically defined CKD in UK Biobank.

Furthermore, T2D risk genes did not generally increase CKD risk in UK Biobank indicating this is a specific effect of IRS-2 disruption. While the mechanistic basis of this association requires elucidation, insulin signalling exerts salutatory effects on podocyte health and function in mice^41–43^ and germline loss of *IRS2* in mice results in smaller kidneys^44^. Both mechanisms could contribute to the adverse effects of PTVs in *IRS2*; podocyte dysfunction and loss is a key early step in many forms of kidney disease including diabetic nephropathy and there is an increasing appreciation that the nephron number at birth - nephron endowment - is an important determinant of kidney health in later life^45,46^, so both of these potential pathogenic mechanisms could be involved. Our demonstration of a causal role for *IRS2* in kidney health provides an important impetus to determine if effects on renal health are mediated by a role of IRS-2 in kidney development and nephrogenesis or by a regulatory role in post-natal renal physiology. If a renoprotective function of IRS-2 in post-natal life exists, then examining the effects of risk factors for renal disease such as diabetes and obesity on IRS-2-mediated signalling could highlight a novel and potentially modifiable mechanism of kidney disease.

Our findings regarding IRS-2 and T2D may be of broad relevance to patients with T2D; it has long been postulated that acquired IRS-2 dysfunction could play a key role in the aetiology of polygenic T2D given its role in two key pathophysiological processes – insulin resistance and pancreatic beta-cell failure^47,48^. Interestingly, a recently described subtype of T2D, severe insulin resistant diabetes (SIRD), presents with reduced eGFR at T2D diagnosis and increased CKD risk that does not seem to be solely related to glycaemic control^49^. Our results highlight impaired IRS-2-signalling as a candidate mechanism in the pathogenesis of this diabetes subtype.

An intriguing finding was the association of the related *UBR2* and *UBR3* with BMI, with the latter also elevating T2D risk in a manner only partially dependent on its effect on BMI. *UBR2* has previously been associated with increased BMI in WES analysis^8^, but this is the first report of damaging mutations in *UBR3* with any cardiometabolic phenotype, to our knowledge. These proteins are both E3-ubiquitin ligases; UBR2 functions as an effector of the N-degron pathway recognising modified N-Terminal amino acids and targeting their host protein for degradation, while UBR3, despite structural homology to UBR2, lacks canonical N-recognin activity^27,28^. Interestingly, while both UBR2 and UBR3 are relatively broadly expressed, UBR3 is enriched in a number of sensory tissues including tongue, ear and olfactory epithelia – which may have relevance to its effects on BMI^17^. Both UBR2 and UBR3 are relatively enriched in expression in skeletal muscle. While the specific effects of these proteins in muscle is unclear, UBR3 may play a non-redundant role in skeletal muscle function as carriers of PTVs in *UBR3* had reductions in grip strength. While our work clearly highlights UBR2 and UBR3 as important regulators of cardiometabolic health, further study exploring their substrates and function are necessary to gain a mechanistic understanding of their effects on BMI and T2D.

There are some potential clinical implications of our findings. Notably, in a phenotypic association scan of potentially relevant traits we observed a strong association between predicted damaging missense mutations in *HNF4A* and reduced circulating SHBG. While it has been noted that HNF4A can activate the SHBG promoter^23^ and a causal relationship between HNF4A and circulating SHBG has been suggested^50^, this is the first such genetic evidence in humans. Pathogenic mutations in *HNF4A* cause a type of monogenic diabetes onset of the young (MODY) - we speculate that individuals with apparent T2D and low SHBG without significant insulin resistance may be enriched for *HNF4A* mutations. We have also identified the first phenotypic consequences of loss of *IRS2* in humans; while damaging mutations in other components of the insulin signalling cascade are reported to cause severe monogenic insulin resistance, the impact of IRS-2 disruption in humans was undocumented. Our work provides an impetus for research-based genetic testing of individuals with T2D and features of severe insulin resistance and in other cases of atypical diabetes^51^, particularly if they also have CKD and/or a monogenic cause is suspected. More generally, it is interesting to speculate that as sample sizes grow, insights from population genetic association studies could increasingly inform clinical intuition regarding the aetiology of diabetes by identification of robustly associated biomarkers in an unbiased manner.

In summary, our study expands the number of genes directly implicated in metabolic health by human gene knockouts, and further illustrates the benefit of genome over exome sequencing for the discovery of rare variants associated with disease.

## Methods

### UK Biobank whole genome sequencing data processing

The whole genome sequencing (WGS) of UK Biobank participants is described in detail in Li et al.^7^ In brief, 490,640 UK Biobank participants were sequenced to an average depth of 32.5X using Illumina NovaSeq 6000 platform. Variants were jointly called using Graphtyper^52^, which resulted in 1,037,556,156 and 101,188,713 high quality (AAscore < 0.5 and <5 duplicate inconsistencies) SNPs and indels respectively.

We further processed the jointly called genotype data in Hail v0.2^53^, where multi-allelic sites were first split and normalized. Variants were then filtered based on low allelic balance (ABHet < 0.175, ABHom < 0.9), low quality-by-depth normalized score (QD < 6), low phred-scaled quality score (QUAL < 10) and high missingness (call rate < 90%). For the analysis in the European ancestry cohort (see below), we further removed variants that failed test for Hardy-Weinbery equilibrium (P<1e-100) within this cohort.

Variants were annotated using Ensembl Variant Effect Predictor (VEP)^54^ v108.2 with the LOFTEE plugin^55^. Combined Annotation-Dependent Depletion (CADD) annotations were based on precomputed CADD^56^ v1.7 annotations for all SNPs and gnomAD v4 indels. REVEL (rare exome variant ensemble learner)^15^ annotations were obtained from the May 3, 2021 release of precomputed REVEL scores for all SNPs. We prioritized the individual consequence for each variant based on severity which was defined by VEP. The protein-truncating variant (PTV) category is the combination of stop-gained, splice acceptor, and splice donor variants. The missense and synonymous variants were adopted directly from VEP. Only the variants on autosomes and the chromosome X, which were within ENSEMBL protein-coding transcripts, were included in our downstream analysis.

### European ancestry definition in UK Biobank WGS

We defined a European-ancestry cohort to be that which most resembled the NFE (non-Finnish European) population as labelled in the gnomAD v3.1 dataset^55^. This NFE group was one of nine ancestry groups labelled in gnomAD, which was based on HGDP and 1000 Genome samples. Variant loadings for 76,399 high-quality informative variants from gnomAD were used to project the first 16 principal components onto all UK Biobank WGS samples. A random forest classifier trained on the nine ancestry labels in gnomAD was then used to calculate probabilities that reflect the similarity between the UK Biobank participant and each of the gnomAD ancestry labels.

### Genome-wide gene burden testing in the UK Biobank

BOLT-LMM^57^ v2.4.1 was used as our primary analytical software to conduct gene burden tests.

To run BOLT-LMM, we first derived a set of genotypes consisting of common (MAF > 0.01) LD-pruned (LD r^2^ < 0.1) variants in individuals with WGS data to build the null model. Pruning was conducted using PLINK2^58^ on a random subset of 50,000 individuals (options in effect: --maf 0.01 --thin-indiv-count 50000 --indep-pairwise 1000kb 0.1).

We adopted the same strategies used in our previous analyses using WES data^9,11^. We generate the dummy genotype files in which each gene-mask combination was represented by a single variant, which were required as the genotype input for BOLT-LMM. We then coded individuals with a qualifying variant within a gene as heterozygous, regardless of the total number of variants they carried in that gene.

We then created the dummy genotypes for the MAF < 0.1% high confidence PTVs as defined by LOFTEE, missense variants with REVEL > 0.5 and missense variants with REVEL > 0.7. After getting all required inputs, BOLT-LMM was used to analyse BMI and T2D using default parameters except for the inclusion of the ’lmmInfOnly’ flag. The covariates included in our analysis are age, age2, sex, age*sex, the first 20 principal components as calculated from all WGS samples, and the WGS-released batch. Different from our previous studies, we included all samples without restricting their ancestries to maximise the sample size. Only samples who withdrew consent or had missing phenotypes and covariates were excluded; filtering resulted in 481,137 and 489,941 samples remaining for BMI and T2D, respectively.

To identify single variants driving a given association within a single gene, we performed a leave-one-out analysis for all identified genes using a generalized linear model in R v4.0.2 by dropping the variants contained in the gene-mask combination one at a time.

As BOLT-LMM use a linear mixed model, we estimated and reported the OR using the generalized linear model in R v4.0.2 for all T2D associated genes.

### Replication in All of Us study

Participants analyzed in this study were selected from the All of Us (AoU) Research Program cohort^33^. The collection of participant information adhered to the AoU Research Program Operational Protocol (https://allofus.nih.gov/sites/default/files/All of Us Research Program Operational Protocol 2022.pdf). Detailed methodologies regarding genotyping, ancestry classification, quality control measures, and the methodology for excluding related participants are thoroughly documented in the AoU Research Program Genomic Research Data Quality Report (https://support.researchallofus.org/hc/en-us/articles/4617899955092-All-of-Us-Genomic-Quality-Report).

We conducted our analysis on short-read whole exome sequencing data (version 7.1), focusing on two phenotypes: BMI and T2D. The analysis encompassed 219,015 unrelated individuals, including 112,526 of European ancestry, 46,414 of African/African American ancestry, 34,865 of American Admixed/ Latino and 25,210 various other ancestries (Supplementary Table 3 for detailed sample size information).

BMI data were derived from the “body mass index (BMI) [Ratio]” metric (Concept Id 3038553) within the “Labs and Measurements” domain. The “Type 2 diabetes mellitus” identifier (Concept Id 201826) in the “Conditions” domain facilitated the identification of T2D cases. For participants with multiple BMI/T2D records, the initial entry was utilized. The participants’ ages were calculated by subtracting the birth year from the timestamp of the earliest record. Among these individuals, 32,462 were identified as T2D cases, and 186,553 served as controls. Only subjects aged over 18 were included in the analyses.

Gene-based burden tests were applied to variants with MAF less than 0.001. These tests were conducted using STAAR (variant-set test for association using annotation information)^59^ implemented in STAARpipeline^60^ (R package version 0.9.7), with covariates adjustments for age, age^2^, sex, age*sex, and the first 16 PCs. The criteria for gene-burden masks followed the methodology of the main UKB analyses.

### UK Biobank whole-exome sequencing processing

To quantify the gain from WGS vs WES in UK Biobank, we compared variant counts between our WGS data with those from the 450K OQFE (original quality functional equivalence) release of the UK Biobank WES data (454,756 individuals total). We processed multi-sample pVCFs using Hail^53^ 0.2, where multi-allelic sites were first split and normalized. Sites were then excluded if they failed the following quality metrics: for SNPs, ABHet < 0.175, QD < 2, QUAL < 30, SOR > 30, FS > 60, MQ < 40, MQRankSum < -12.5 and ReadPosRankSum < -8; for indels: ABHet < 0.175, QD < 2, QUAL < 30, FS > 200 and ReadPosRankSum < -20, resulting in 23,273,514 variants available for analysis. Individuals with high heterozygosity rates, discordant WES genotypes compared to array and discordant reported versus genetic sex were removed, resulting in 453,931 individuals. Variants were annotated using the identical VEP pipeline, LOFTEE, CADD and REVEL annotations as described for WGS.

### Phenotypic association scan of identified BMI and T2D associated genes in UKBB

We ran association tests between each identified genes carriers and a list of representative phenotypes (full list can be found in Supplementary Table 7 and 8) available in the UK Biobank using R v4.0.2 including the same covariates we used in our genome-wide gene burden tests. We also extracted the phenotypic associations with *P*<0.05 for all genes we identified in our analysis from AstraZeneca PheWAS Portal^61^ (version: UK Biobank 470K WES v5, Supplementary Table 9 and 10).

### BMI and T2D GWAS lookup

Identified genes were queried for proximal BMI and T2D GWAS signals, using data from the largest published GWAS meta-analyses. For BMI, we used data from the GIANT consortium^62^, which includes data on up to 806,834 individuals. For T2D, we used data from the DIAGRAM consortium^63^, which included up to 428,452 T2D cases and 2,107,149 controls.

For each of these GWAS, we performed signal selection and prioritised causal GWAS genes using the “GWAS to Genes ”pipeline as described elsewhere^18^. The previously identified genes were annotated if their start or end sites were within 500kb up- or downstream of GWAS signals in the two meta-analyses, using the NCBI RefSeq gene map for GRCh37, and overlayed with further supporting functional dataset information. For further details about the specific application of this method, see Kentistou et al.^18^

## Code availability

All analyses were performed used publicly available softwares. No custom code was developed.

## Data Accessibility

The UK Biobank phenotype, whole-genome and whole-exome sequencing data described here are publicly available to registered researchers through the UKB data access protocol. Information about registration for access to the data is available at: https://www.ukbiobank.ac.uk/enable-your-research/apply-for-access. Data for this study were obtained under Resource Application Numbers: 20361 and 68574.

The All of Us phenotype and whole-genome sequencing data described here are available to registered researchers through the All of Us data access protocol. Information about registration for access to the data is available at: https://www.researchallofus.org/register/ .

## Data Availability

The UK Biobank/All of Us cohort phenotype and WGS data described here are publicly available to registered researchers through the UK Biobank/All of Us cohort data access protocol.

